# Excellent repigmentation was observed in the treatment of refractory vitiligo with autologous cultured epithelium grafting: a real-world retrospective cohort study

**DOI:** 10.1101/2022.12.18.22283394

**Authors:** Jian Li, Xuanhao Zeng, Shujun Chen, Luyan Tang, Qi Zhang, Minzi Lv, Taro Uyama, Fuyue Wu, Weiling Lian, Jinqi Wang, Haozhen Lv, Yating Liu, Jinfeng Wu, Jinhua Xu

**Affiliations:** Department of Dermatology, Huashan Hospital of Fudan University, Shanghai, China; Centre of evidence medicine, Fudan Univerisity, Shanghai, China; ReMed Regenerative MedicineClinical Application Institute, Shanghai, China

## Abstract

Surgical intervention is considered as the mainstream therapy for refractory vitiligo. In this study, we developed a modified autologous cultured epithelial grafting (ACEG) technique for the surgical treatment of vitiligo. A total of 726 patients with vitiligo treated with ACEG were enrolled from January 2015 to June 2019 in China. Patient characteristics, such as sex, age, clinical type, lesion sites, course of the disease, and disease stable period, were recorded. In 2118 skin lesions from 726 patients who received ACEG, total efficacy rate was 82.81% (1754/2118).

However, the repigmentation rate of the ACEG was 64.87%, which was higher than that of conventional surgical interventions (52.69%). Patients with segmental vitiligo, skin lesions in the lower limbs, aged 18 years or below, and a stable period of over 3 years might have a good response to ACEG. Single-cell RNA sequencing was performed to observe different cell compositions in the skin before and after ACEG. The number of melanocytes increased by 50% after transplantation. In addition, there was a significant increase in hair follicle outer root sheath-derived keratinocytes in ACEG, and the numbers of these cells in the repigmentation sites 1 year after ACEG were still higher than those in the skin lesions. Therefore, ACEG is a promising therapeutic agent for refractory vitiligo. Age, clinical type, lesion site, and lesion stable period before surgery have significant impacts on repigmentation in ACEG. ACEG can increase the number of melanocytes and KRT6C+ keratinocytes in skin lesions, thereby restoring a skin microenvironment suitable for melanocyte survival.

**One sentence summary:** Autologous cultured epithelial grafting (ACEG) technique is a promising therapy for refractory vitiligo.

## INTRODUCTION

Vitiligo is a skin disease characterized by acquired depigmentation(*1*). Although vitiligo is not fatal, it significantly affects patients’ quality of life (QOL) through psychological burden and social exclusion(*2*). Fifty-seven percent of patients have reported that vitiligo moderately or severely affect their QOL(*3*). Conventional treatments for vitiligo include: phototherapy, topical corticosteroids, and calcineurin inhibitors. However, these treatment options only provide partial repigmentation in many patients(*4*).

Surgical interventions are mainstream therapies for refractory vitiligo that are resistant to conventional approaches. Surgical techniques for vitiligo include tissue and cellular grafting (*5*). Tissue grafting includes: punch grafting, suction blister epidermal grafting (SBEG), and thin skin grafting(*6, 7*). There are three types of cellular grafting: cultured epidermal cell suspension (CECS), noncultured epidermal cell suspension (NCES), and noncultured follicular cell suspension (NCFC)(*8*). In a recent systematic review and meta-analysis that included 117 unique studies and 8776 unique patients, the rates of repigmentation above 90% and 50% after a single session of all surgical interventions were 52.69% and 81.01%, respectively(*9*). However, conventional surgical interventions have several limitations. For example, owing to the limited size of the normal epidermis, tissue grafting is not suitable for large areas of vitiligo, whereas cellular grafting is not easily fixed in skin lesions of vitiligo, and usually causes uneven repigmentation in special parts of the body, such as the nose and lip(*10*).

In 1975, Green et al. first cultivated human keratinocytes on a feeder layer of lethally irritated 3T3-J2 mouse embryonic fibroblasts, and then obtained cultured epidermal sheets (*11*). The advantage of this technique is that it can obtain a large epidermal sheet by consuming a relatively small epidermis. In 2013, Kumagai et al. adopted Green’s technique for the treatment of 27 vitiligo patients, and reported good repigmentation(*12*). However, this technique also has several limitations. Human keratinocytes can be cultivated on a feeder layer of mouse fibroblasts in fetal bovine serum-containing medium, which might contain potentially harmful agents, such as bovine spongiform encephalopathy and murine viruses(*13*).

To overcome these limitations, we developed a modified autologous cultured epithelial grafting (ACEG) technique under serum-free and feeder-free conditions. We have applied this technique in treating over 1000 patients with vitiligo since 2015. To obtain substantial evidence regarding the efficacy and safety of this technique for refractory vitiligo, we conducted a real-world study with a large-scale sample size.

To explore the mechanism of repigmentation after ACEG treatment, single-cell RNA sequencing (scRNA-seq) was performed to observe the differences in cell composition in the skin of patients with vitiligo before and after ACEG treatment.

## RESULTS

### Patient characteristics and therapeutic effects of ACEG

This retrospective study included 790 patients with vitiligo who were treated with ACEG. Patients who lacked data or had poor photo quality, as well as those who could not be contacted, were excluded. Furthermore, 726 patients were included, with a total of 2118 skin lesions treated with ACEG, including 347 men (47.8%) and 379 women (52.2%) (Fig.1). The average age was 23.56 ± 9.5 years old, with the youngest being 6 years, and the oldest being 66 years. Among the clinical types, 460 (63.36%) were of the segmental type. The average course of disease was 8.43±7.35 years. Before the operation, the stable period for vitiligo ranged from 6 months to 40 years, and the longest stable period was 45 years. The largest single lesion area was 394 cm^2^. Among the 2118 pieces of skin lesions, the repigmentation of 1374 pieces (64.87%) was excellent, and 380 pieces (17.94%) were good, with a total effective rate of 82.81% (Fig.2).

**Fig 1.**
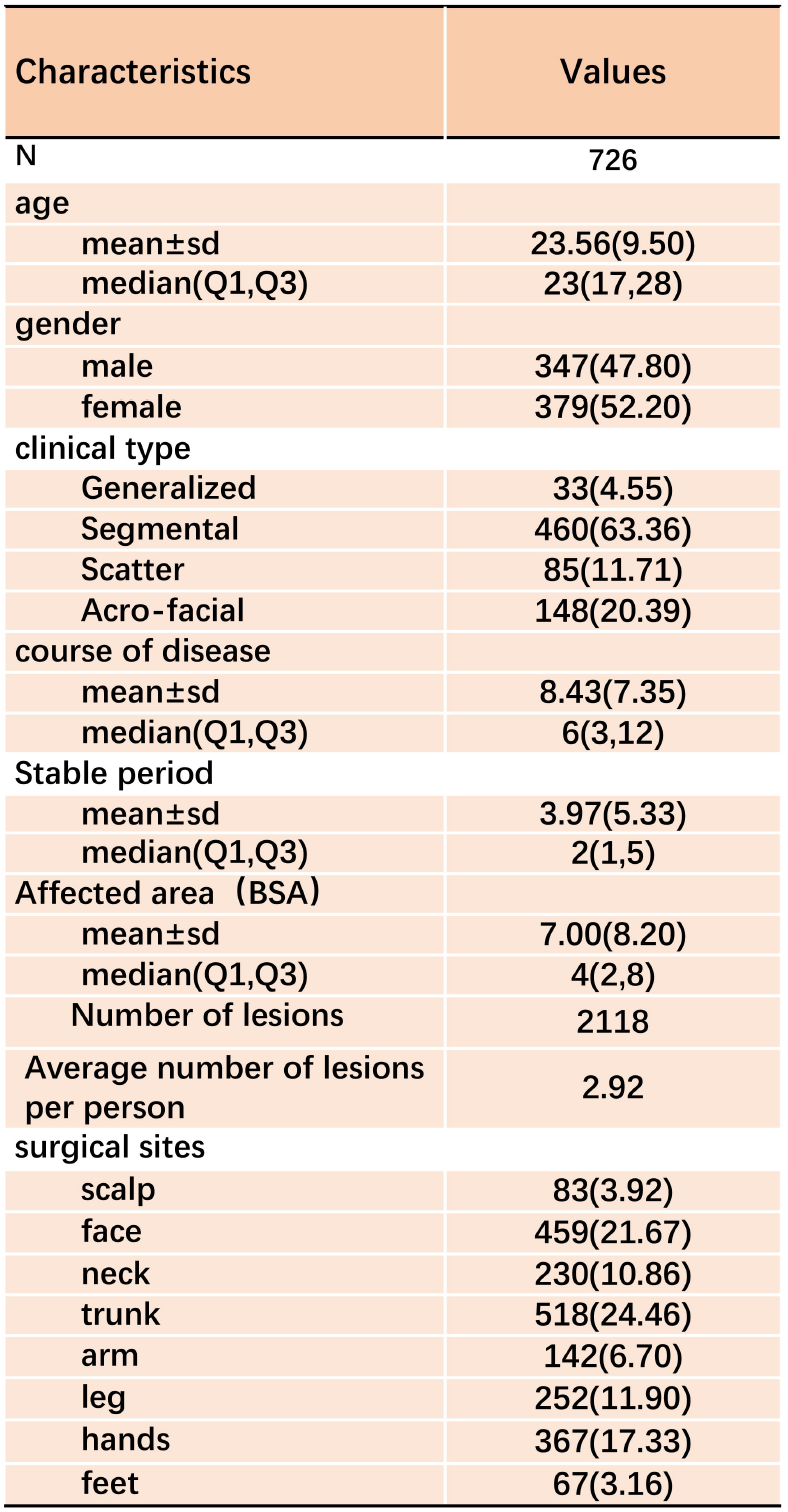
Patient characteristics.

**Fig 2.**
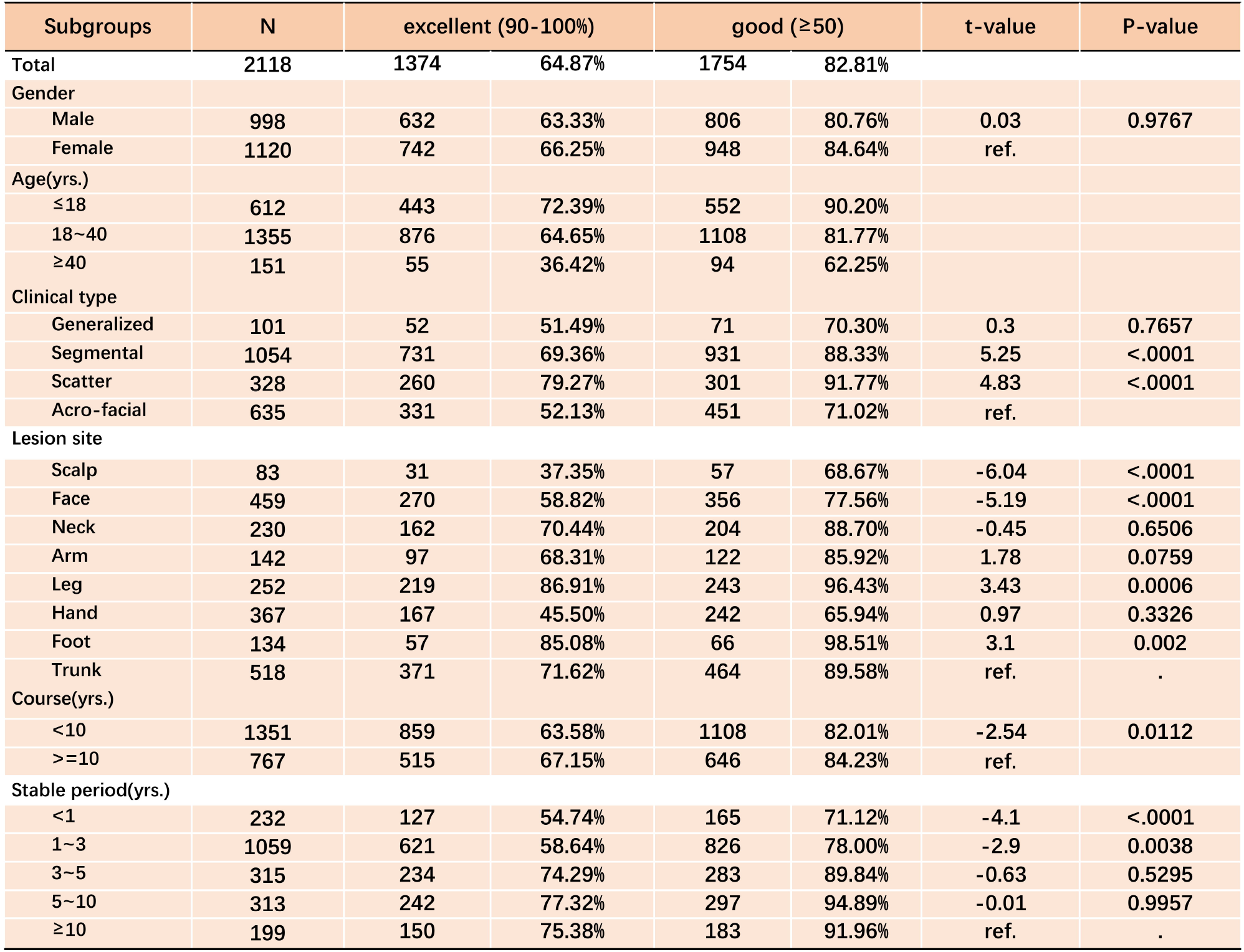
Therapeutic effects of ACEG and correlation analysis with different clinical factors. The correlation analysis of therapeutic effects with gender, age, disease type, lesion site, course of disease, and stable period.

### Correlation analysis of therapeutic effects with gender, age, disease type, lesion site, course of disease, and stable period

Among the 2118 skin lesions, 998 were from men and 1120 from women (Fig.2). There was no difference in the efficacy rate between men and women (80.76% versus 84.64%). Age has a significant influence on patient repigmentation. The effective rate of patients aged 18 years and below was 90.20%, while that of patients aged over 40 years was only 62.25%. Different types of vitiligo have different rates of repigmentation. The effective rate of segmental vitiligo and scatter vitiligo (88.33%, 91.77%) was significantly higher than that of generalized vitiligo and acral vitiligo (70.3%, 71.02%). Different lesion sites affected the repigmentation rate in the ACEG. The effective rate in the foot and lower limbs was the highest (98.51% and 96.43%, respectively), while it was only 77.56% in the face. Among all lesion sites, the effective rates on the scalp and hand were the lowest (68.67% and 65.94%, respectively). There was no difference in the effective rate between over and less than 10 years of the disease course (82.01% versus 84.23%). However, the stable period of the disease had a marked impact on the repigmentation rate in ACEG. The effective rate of patients who were stable within one year was significantly lower than that of patients who were stable for more than three years (71.12% vs. 89.84%), and the effective rate of patients who were stable for 5–10 years was the highest (94.89%).

### Adverse events

In a total of 726 patients, 10 had scar hyperplasia, involving 24 skin lesions, especially in the chest, back, and upper limbs. Two patients developed infection in the lower limbs. One patient with a severe case was hospitalized, and improved three days after cephalosporin injection. Nine patients reported new leukoplakia after ACEG.

### Comparison of skin cell composition before and after ACEG treatment

scRNA-seq analysis demonstrated that the cell constitution between the lesion and repigmented skin was similar. More than 90% of the clusters in these two samples had a similar constitution, which indicated that ACEG cannot significantly change the cell composition of the epidermis (**S Fig.2&3**). As expected, the proportion of melanocytes showed a 50% increase in skin repigmentation compared with skin lesions. In keratinocyte clusters, the repigmentation sample showed a remarkable increase in the proportion of KRT6C+ keratinocytes compared to the sample from the lesions (2% versus 0.09%), which characterized by high expression of hair follicle outer root sheath-derived cell-related genes, such as KRT6C, KRT16, and KRT17(*14*), which may support melanocyte growth(*15*).We Also, cells secrete proteins that promote melanocyte proliferation and differentiation, including EDN1, FGF7, HGF and Laminin, are significant up-regulated in skinsheet. One year after ACEG, the proportions of these cells were significantly higher in the repigmented skin than in the skin lesions, which indicated that these cells might play important roles in repigmentation (Fig.4). These results were further verified by immunofluorescence (S Fig.4).

**Fig 3.**
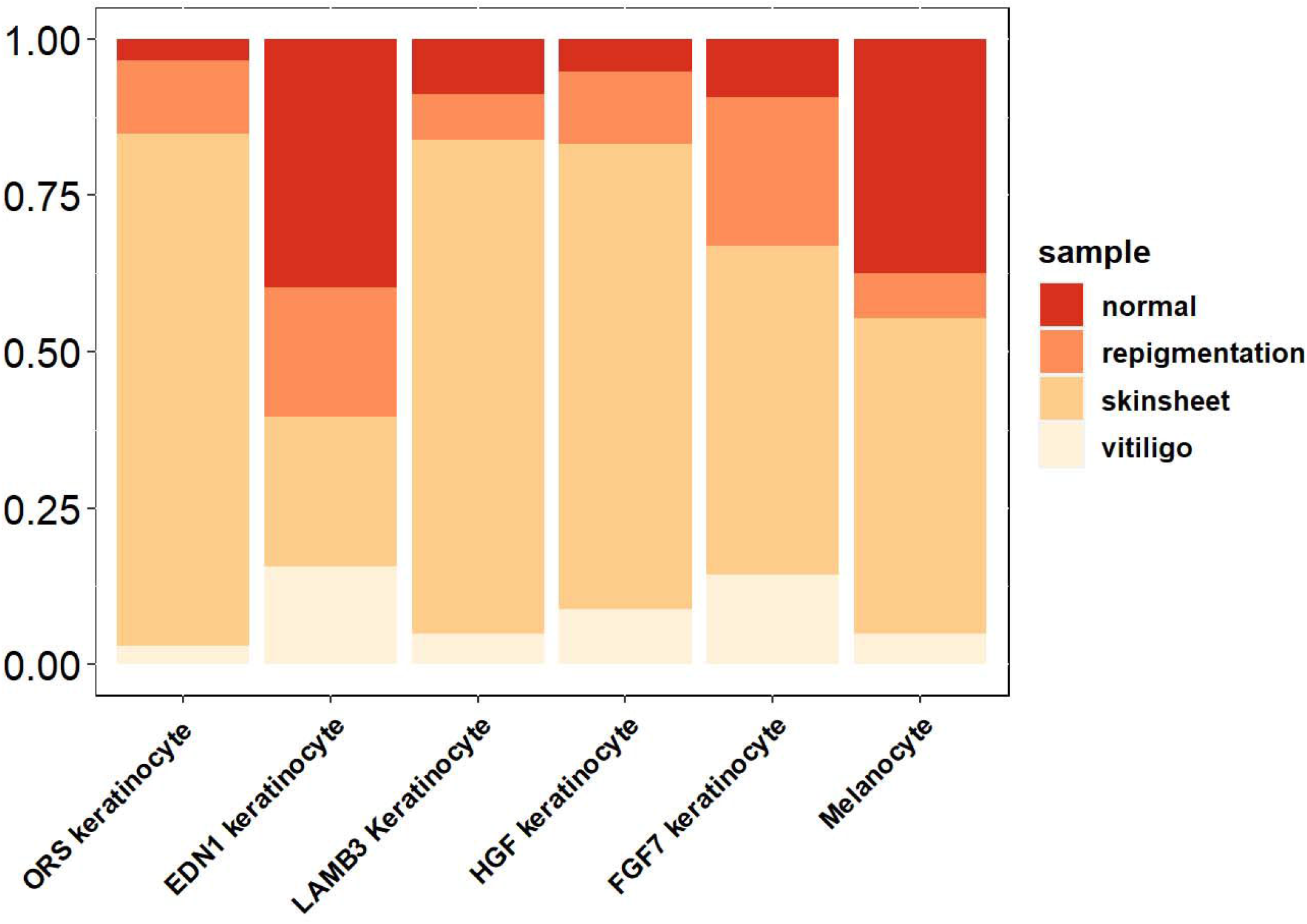
sc-RNA seq analysis of vitiligo skin samples. The proportion of different cell clusters in four skin samples. normal, normal skin sample from the groin; vitiligo, depigment skin lesion from the patient; repigmentation, same site repigment one year after the ACEG treatment; skinsheet, ACE made from normal skin.

**Fig 4.**
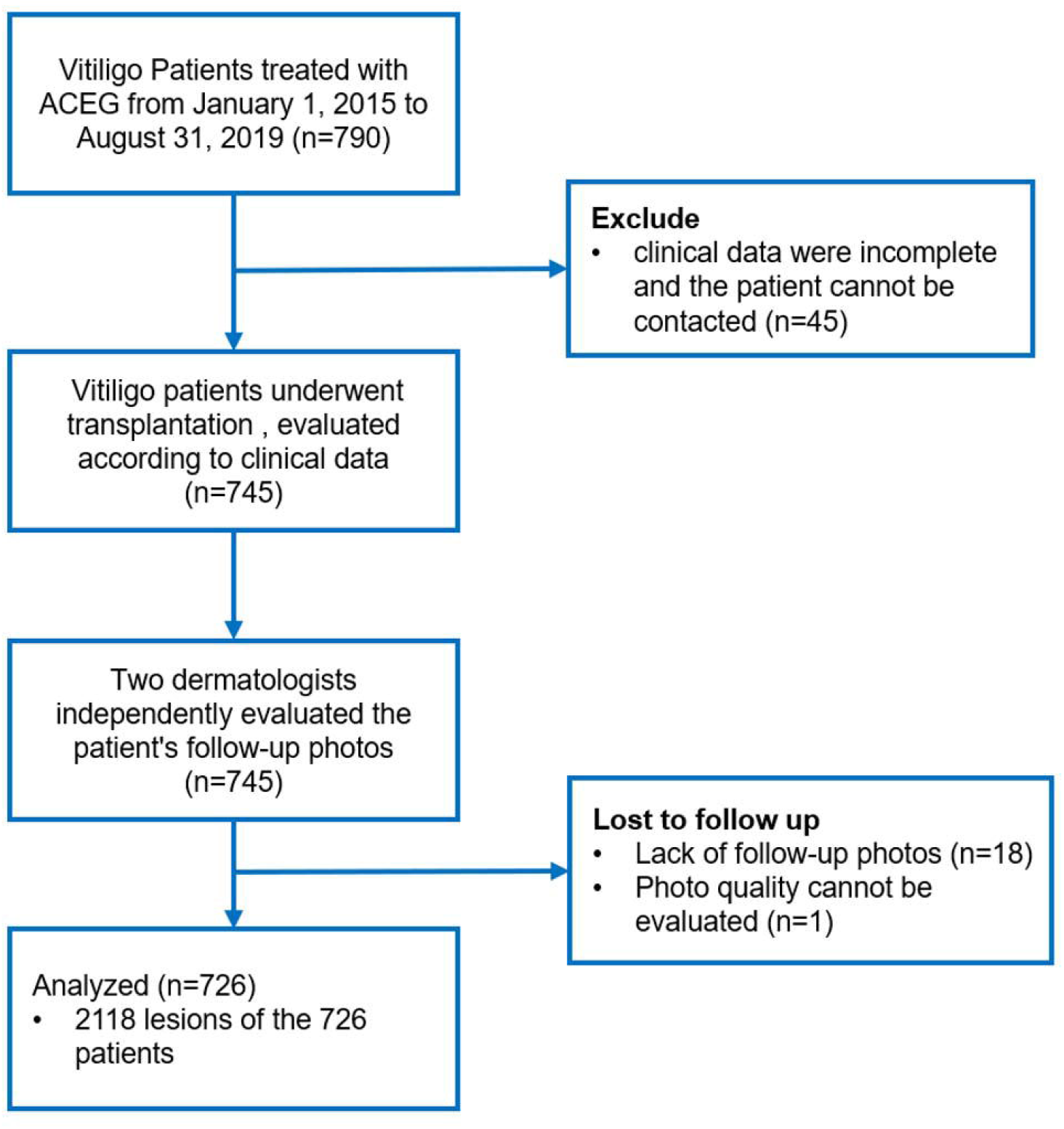
Trial profile.

## DISCUSSION

Conventional ACEG cultivates human keratinocytes with mouse fibroblast feeders in fetal bovine serum-containing medium, which contains animal ingredients and might have potentially harmful effects (*13, 16*). To avoid the possible disadvantages of the conventional ACEG, we developed a modified ACEG method under serum-free and feeder-free conditions. Our previous study showed no significant difference in the rate of repigmentation between conventional and modified ACEG(*17*). We have performed a modified ACEG for over 1000 patients with vitiligo since 2015. To obtain substantial evidence regarding the efficacy and safety of this technique for the treatment of refractory vitiligo, we conducted a real-world study with large-scale samples.

The efficacy rate of surgical intervention for vitiligo was previously determined according to the efficacy of a single patient(*18*). However, patients with vitiligo usually have more than one lesion that should be treated with ACEG. Different lesions in the same patient may have different effects. In this study, we analyzed the efficacy of ACEG in a single lesion instead of a single patient, and found that the total efficacy was similar to that of conventional surgical interventions (82.81% vs. 81.01%). Next, we demonstrated that the rate of repigmentation over 90% was 64.87%, which was higher than that of conventional surgical interventions (64.87% vs. 52.69%). Although there was no difference in the total effective rate between ACEG and conventional surgical interventions, ACEG might have advantages over conventional surgical interventions in terms of an excellent repigmentation rate.

We found that age significantly influenced repigmentation. For example, the effective rate in patients aged 18 years and below was 90.2%, while the effective rate in patients over 40 years old was only 62.25%. Young patients receiving ACEG might benefit more than older patients. Grafted stem cells should be young and devoid of senescent defects for regenerative purposes(*19*). It is highly possible that the cells from the donor epidermis of vitiligo patients aged over 40 years were senescent compared to those in vitiligo patients aged 18 years and below. Furthermore, we observed that different clinical types of vitiligo have different repigmentation rates. The efficacy rates of ACEG in segmental vitiligo and scatter vitiligo (88.33%, 91.77%) were considerably higher than those in generalized vitiligo and acral vitiligo (70.3%, 71.02%). This result regarding the influence of vitiligo type on the efficacy of ACEG is in line with that of conventional surgical techniques (*9, 20-22*). The location of skin lesions affects repigmentation(*20, 23, 24*).The effective rates in the foot and lower limbs were the highest (98.51% and 96.43%, respectively), while the effective rates in the scalp and finger were the lowest (68.67% and 65.94%, respectively). The finger frequently moves, and the blood supply to the finger is relatively poor. Although the blood supply to the scalp is abundant, the epidermis sheet is not easily fixed in the scalp because of hair. These factors may affect the efficacy of ACEG. The stable period of vitiligo before ACEG treatment had a significant impact on remission rate. The effective treatment rate of patients who were stable within one year (71.12%) was considerably lower than that of patients who were stable for more than three years (89.84%). A longer stable period of skin lesions may create a more suitable microenvironment for melanocyte growth and repigmentation.

The classic mechanism of vitiligo with surgical intervention is the reconstruction of the affected melanocyte reservoir after transplantation(*25*). Our sc-RNA-seq analysis demonstrated that the number of melanocytes after ACEG increased by 50%. In addition, it has become a consensus that keratinocytes in vitiligo lesion have abnormal state, morphology, and function(*26*). They do not normally secrete cytokines, such as SCF and EDN1, which are important for the growth and function of melanocytes (*27*). The abnormal state of keratinocytes in vitiligo is difficult to recover after formation, which might cause failure of repigmentation in vitiligo(*14*). In this study, we found that there were many hair follicle outer root sheath-derived cells in ACE, and the number of HFORS in the repair sites 1 year after ACEG was still higher than that in the lesion skin, which may be related to wound repair (*28*).

The limitations of our study are related to its observational nature, and we cannot exclude bias owing to uncontrolled (residual) confounding. First, the indicators observed in this study were limited, and some factors affecting the study were not considered in the analysis, such as BMI and previous treatment regimens received by patients. Second, there was no direct comparison with other transplantation methods in this study, because blister transplantation is not suitable for the treatment of a large area of patients. In addition, other transplantation methods have not been approved by the Chinese Food and Drug Administration in China; thus, a good comparison in China cannot be obtained. Although based on literature review our efficiency is similar to that of other transplantation methods, the actual efficiency may be biased.

Owing to the high technical difficulty of the skin transplantation treatment method used in this study, most hospitals in China are not currently qualified to conduct this project; therefore, this study is a single-center study. Furthermore, we are trying to promote our treatment method and strive to conduct multicenter studies in the future. Meanwhile, the current price of tissue engineering transplantation is relatively high, and most patients who can afford this cost have a good economic foundation.

In particular, we confirmed the efficacy and safety of ACEG in a real-world study, and found that ACEG is effective and safe for treating refractory vitiligo. Patients with segmental vitiligo in the lower limbs, aged 18 years or below, and a stable period of > 3 years might be more suitable for ACEG. ACEG may reverse the abnormal keratinocyte state in the lesion area of vitiligo, thereby restoring the skin microenvironment suitable for melanocyte survival and repigmentation. Thus, ACEG is a promising therapeutic agent for refractory vitiligo.

## MATERIALS AND METHODS

### Trial design

We conducted a single-center retrospective study with a large sample size. This study was approved by the Ethics Committee of Huashan Hospital, Fudan University (KY2020-565), and registered with Chictr. org. cn (ChiCTR2100051405).

### Participants

The data of vitiligo patients treated with ACEG were collected from January 2015 to June 2019, and these patients completed 6 months of clinical follow-up after transplantation (**Fig. 5**). In total, 790 patients were included in this study. Patient characteristics, such as sex, age, clinical type, lesion sites, course of the disease, and disease stable period, were recorded.

Efficacy was determined by two dermatologists based on the follow-up photographs. Repigmentation was evaluated on the 6th month after surgery, and was graded as excellent (90-100%), good (≥50% and <90%), fair (<50% and ≥20%), and poor (<20%). Repigmentation of over 50% was considered effective. Adverse reactions were also recorded.

### Autologous Cultured Epithelium

The donor skin was obtained as a full-thickness skin biopsy specimen (3-6 cm^2^) from a normally pigmented site, usually the groin. Skin tissue was digested with trypsin after thorough sterilization. Trypsinized epidermal cells were collected and cultured in clinical keratinocyte medium (CKM), that is, a xeno-free chemically defined medium containing IGF-1, KGF, bFGF, α-MSH and 0.1 mM Ca^2+^. Clinically approved antibiotics gentamicin and erythromycin were added to prevent contamination.

After passaging, epidermal cells were cultured in CKM for three days, and then switched to keratinocyte growth medium KGM (ReMed), which contains chemically defined ingredients and 1.4 mM Ca^2+^ to promote the maturation of keratinocytes. After seven to ten days, the epidermal cells formed an autologous cultured epithelium (ACE). This epidermal sheet has a normal proportion of melanocytes, as proven by DOPA staining, and is ready for transplantation.

### Transplantation of ACE

Transplantation of ACE was performed as described in our previous published paper(*17*)

### Sc-RNA seq Analysis

For scRNA-seq analysis, we enrolled one patient who received ACEG treatment(**S Fig.1**). This study was approved by the Ethics Committee of the Huashan Hospital, Fudan University (KY2020-1137). Four different skin samples were obtained from the same patient: 1) donor skin: skin sample from the donor site, which was used for the generation of skin sheets; 2) skin sheets: autologous cultured epithelium derived from the donor skin; 3) Lesion skin: skin sample from skin lesions before transplantation; and 4) Repigmentation Skin: skin sample from the ACEG-treated site 1 year after surgery.

The single-cell suspension was prepared and loaded onto the 10X Chromium Single Cell Platform (10X Genomics) at a concentration of 1000 cells/μL (Single Cell 3 library and Gel Bead Kit v.3), as described in the manufacturer’s protocol. For each sample, 5000 cells were captured. Generation of gel beads in emulsion (GEMs), barcoding, GEM-RT cleanup, complementary DNA amplification, and library construction were performed according to the manufacturer’s protocol. A Qubit was used for library quantification prior to pooling. The final library pool was sequenced on an Illumina Novaseq 6000 instrument using 150-base-paired-end reads. Scrublet (version Python 3.7.3) and Cellranger (version 4.0.0) were used to generate clean data. Seurat (3.2.0) was used to analyze the subcluster and marker genes. The trajectory was analyzed using Monocle2 (2.12.0). Gene Ontology (GO) enrichment and KEGG analyses of differentially expressed genes were performed using the clusterProfiler R package with gene length bias correction. Ingenuity Pathway Analysis (IPA) analysis was conducted at Fudan University.

### Statistical analysis

Continuous variables were presented as mean (standard deviation) or median (interquartile range), and categorical variables were described as frequencies and percentages. The 95% confidence intervals (CIs) of the repigmentation and efficiency rates were calculated using the Wald asymptotic confidence limits. The results of the subgroups are presented as forest plots. The pre-specified subgroups included sex (female and male), age (≤18, 18–40, and≥40 years), clinical type (generalized, segmental, scatter, and acro-facial), lesion type (scalp, face, neck, trunk, arm, leg, hand, and foot), course of the disease (<10 and > ≥ 10), and disease stable period (<1, 1–3, 3–5, 5–10, and≥10).

The generalized linear mixed model was used to explore the potential association between the clinical characteristics and repigmentation rate. Models were constructed with age, sex, clinical type, course, and lesion site as fixed factors, and subject (intercept) as random factors.

All hypothesis tests were two-sided, and p values <0.05 were considered statistically significant. All data were analyzed using SAS software (version 9.4) and R software (version 4.1.1).

## Data Availability

All data produced in the present study are available upon reasonable request to the authors

https://www.ncbi.nlm.nih.gov/

## Abbreviation and acronym list

ACEG: autologous cultured epithelium grafting
QOL: quality of life
SBEG: suction blister epidermal grafting
CECS: cultured epidermal cell suspension
NCES: noncultured epidermal cell suspension
NCFC: noncultured follicular cell suspension
scRNA-seq: single-cell RNA sequencing
CKM: clinical keratinocyte medium
KGM: keratinocyte growth medium
ACE: autologous cultured epithelium
CIs: confidence intervals
KRT: keratin
HGF: Hepatocyte growth factor
FGF: Fibroblast Growth Factor
SCF: Stem Cell Factor
EDN: Endothelin
HFORS: hair follicle outer root sheath
BMI: Body Mass Index
LAMB3: Laminin subunit beta-3

## SUPPLEMENT FIGURE LEGENDS

**S Fig.1.**
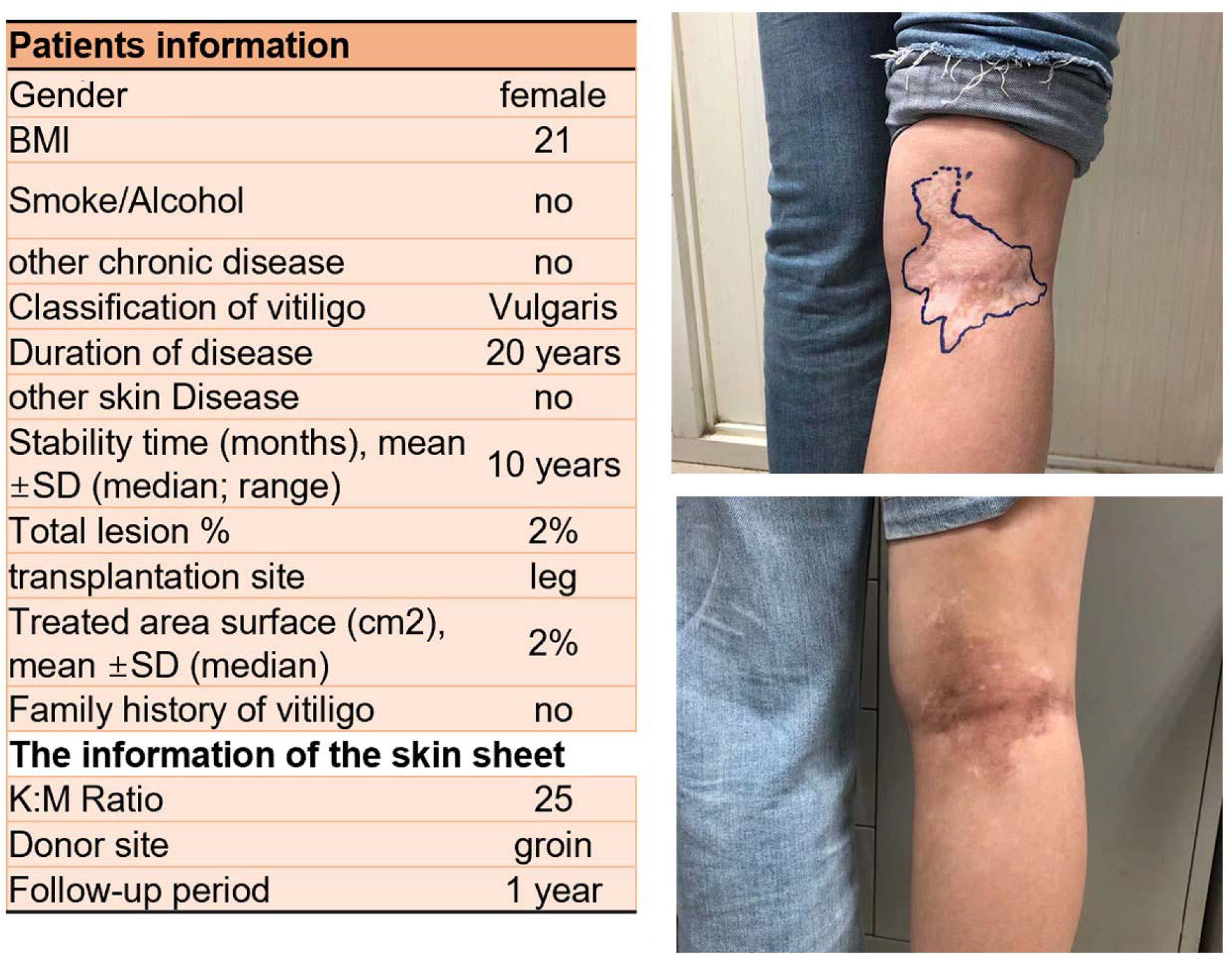
The basic information of vitiligo patient in scRNA-Seq. One year after the ACEG treatment, the lesion is recovered

**S Fig.2.**
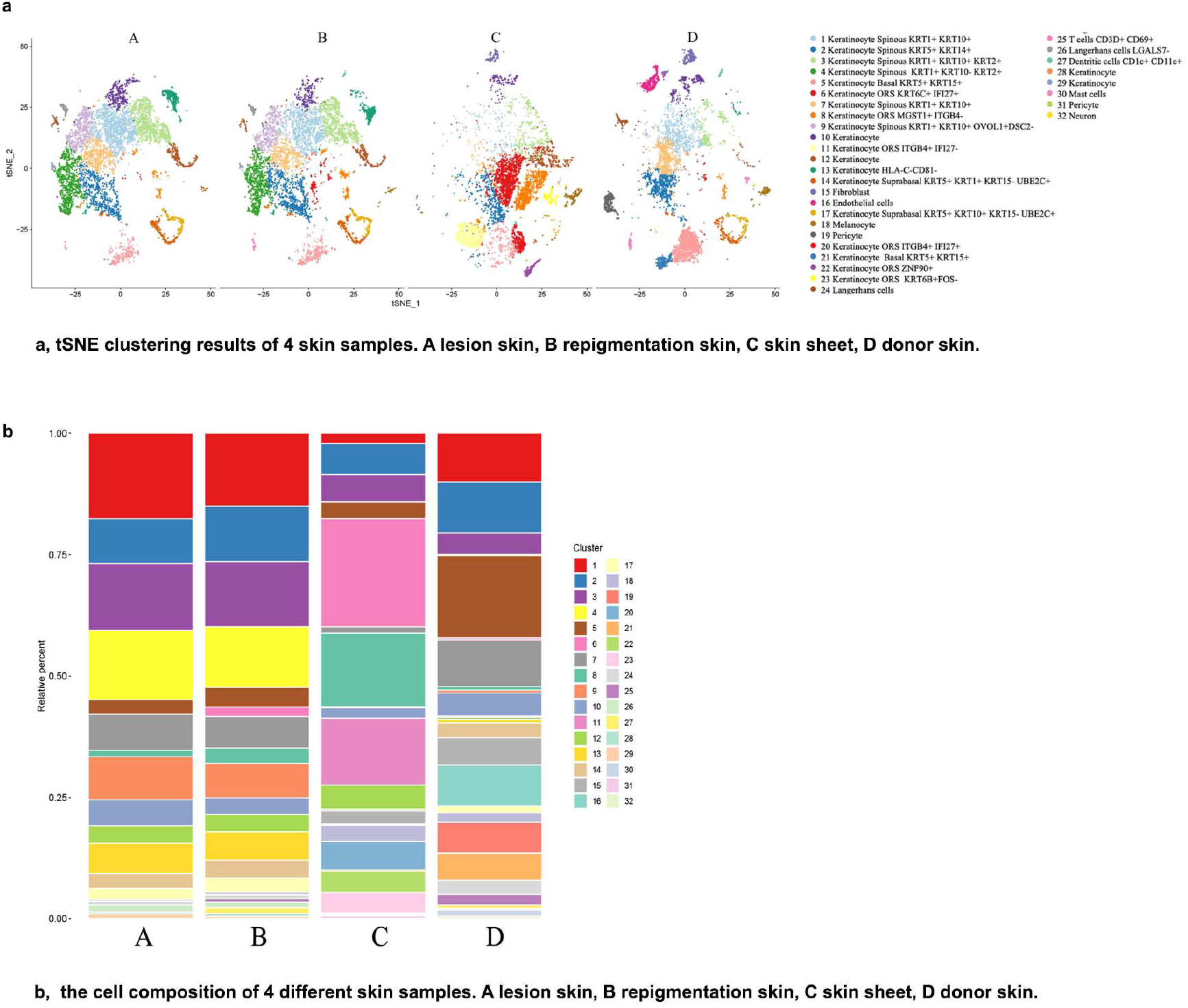
scRNA-seq results. **a**, tSNE clustering results of 4 skin samples. A lesion skin, B repigmentation skin, C skin sheet, D donor skin.b, the cell composition of 4 different skin samples. A lesion skin, B repigmentation skin, C skin sheet, D donor skin

**S Fig.3.**
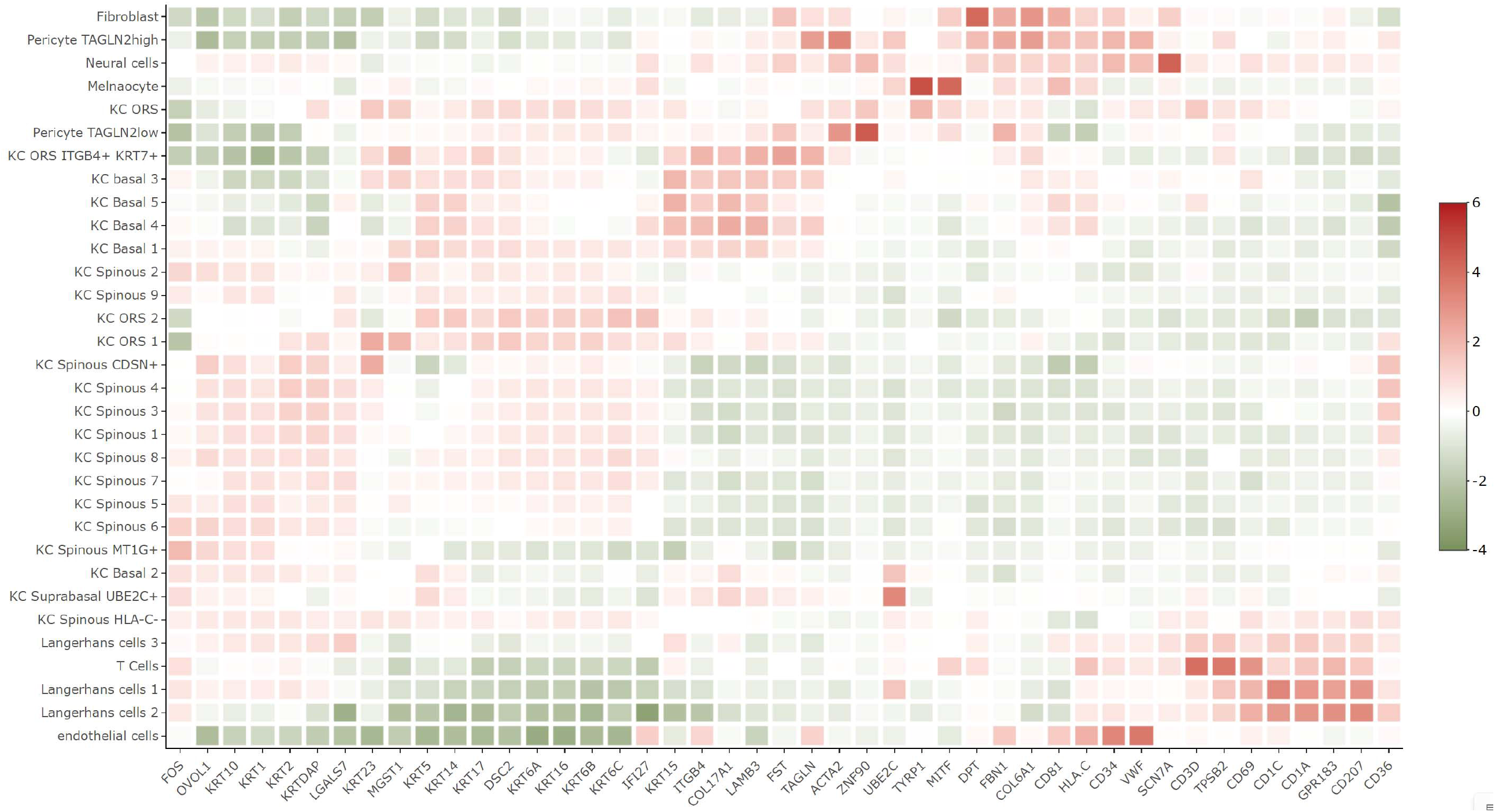
heatmap of major marker genes in scRNA-seq.

**S Fig.4.**
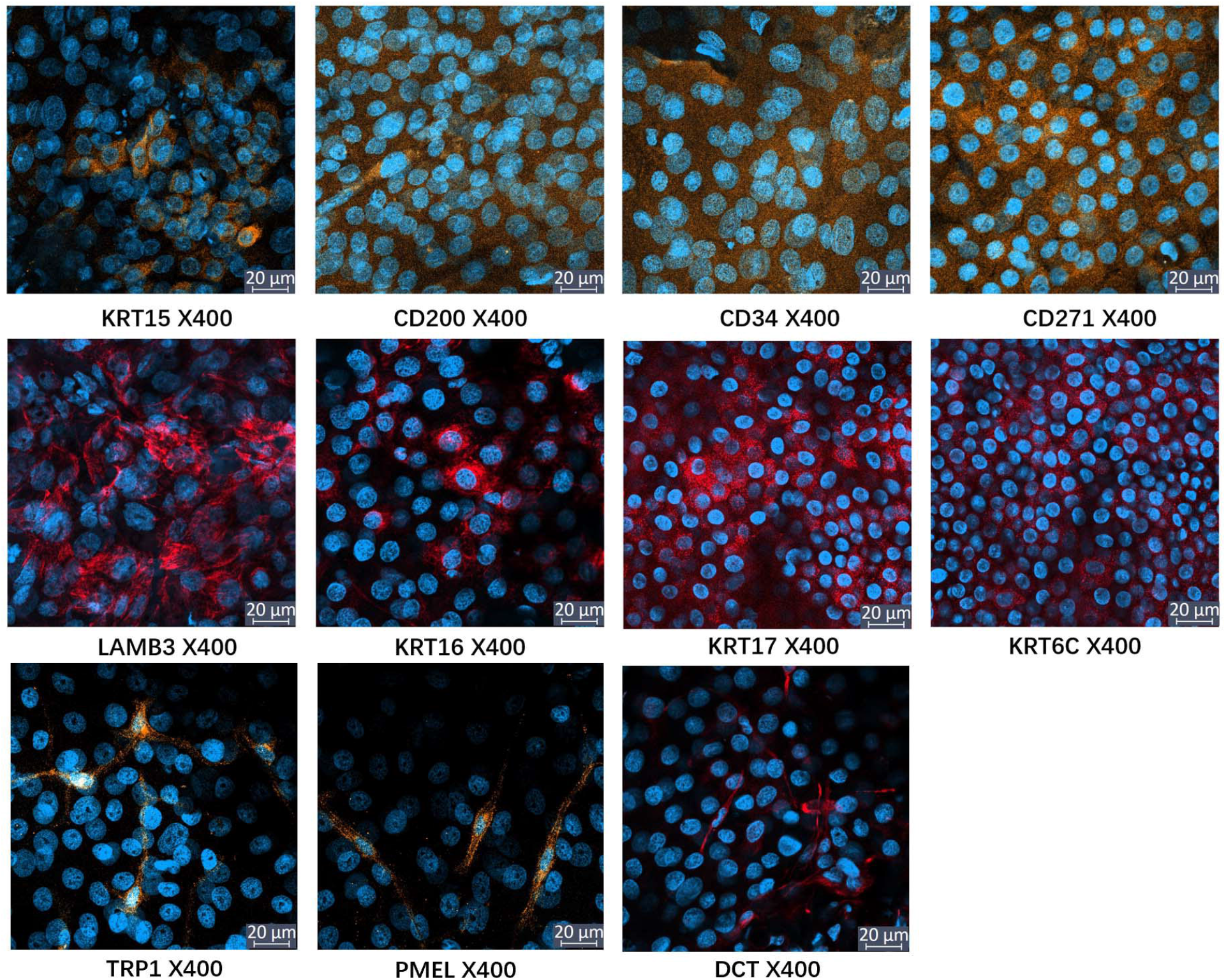
Immunostain results of major marker genes in scRNA-seq.

## Author contributions

Conceptualization: JX, JL

Methodology: JX, XZ, JL, SC, LT, JW, JLS, EH, TU, FW

Investigation: JL, SC, XZ, WL, HL, JW

Visualization: XZ, JL, ML

Funding acquisition: JL, JX

Project administration: JX, JW, XZ

Supervision: JX, JW, JL

Writing – original draft: JL, JW, XZ, LT, QZ

Writing – review & editing: JX, JW

## Data and materials availability

The gene sequencing data in this article has been deposited to https://www.ncbi.nlm.nih.gov/ as SRR18079681, SRR18079680, SRR18079679, SRR18079678. All data, codes, and materials in the analysis can be provided to any researcher for purposes of reproducing or extending the analysis by contact with the correspondence author too. All data, codes, and materials used in the analysis must be available in some form to any researcher for purposes of reproducing or extending the analysis. Include a note explaining any restrictions on materials, such as materials transfer agreements (MTAs). Note accession numbers to any data relating to the paper and deposited in a public database; include a brief description of the dataset or model with the number. If all data are in the paper and supplementary materials, include the sentence “All data are available in the main text or the supplementary materials.”

## Research ethics and patient consent

IRB approval status: Reviewed and approved by Huashan hospital Fudan University; approval KY2020-565. Registered with Chictr.org.cn (ChiCTR2100051405).

